# Neuroinflammatory Disorders of the Central Nervous System Associated with Monkeypox Virus Infection: A Systematic Review and Call to Action

**DOI:** 10.1101/2024.09.14.24313675

**Authors:** Shramana Deb, Ritwick Mondal, Purbita Sen, Dipanjan Chowdhury, Shramana Sarkar, Granthik Banerjee, Vramanti Sarkar, Anjan Chowdhury, Julián Benito-León

## Abstract

**Background:** The Monkeypox virus, a zoonotic pathogen of the Orthopoxvirus genus, has shown a marked global spread, resulting in cases of neuroinflammatory disorders. This systematic review aims to summarize the central nervous system manifestations linked to monkeypox virus infection.

**Methods:** We conducted a systematic review according to PRISMA guidelines. Databases such as PubMed, EMBASE, Cochrane Library, and Web of Science were searched up to September 2024. Inclusion criteria focused on monkeypox virus-positive patients with confirmed central nervous system inflammatory disorders, including encephalitis, meningitis, myelitis, and related neuroimmune diseases.

**Results:** From 770 screened articles, 38 studies were included. Of the 20 reported monkeypox virus-infected cases with central nervous system involvement, the most common manifestations were encephalitis (40%), encephalomyelitis (30%), and meningoencephalitis (10%). Additional rarer presentations included Guillain-Barré syndrome (5%) and transverse myelitis (5%). The mean age of affected individuals was 27.9±13.69 years, and 75% were male. Fever, fatigue, and myalgia were the predominant systemic symptoms, with neurological complications often appearing within 4.6±2.5 days post-infection. Diagnoses were confirmed by quantitative reverse transcription polymerase chain reaction, magnetic resonance imaging, and cerebrospinal fluid analysis. Treatment strategies varied, with antivirals (e.g., tecovirimat) and immunomodulatory therapies (e.g., methylprednisolone, immunoglobulins, and rituximab) utilized.

**Conclusions:** Monkeypox virus infection is associated with severe neuroinflammatory disorders, with varying outcomes from complete recovery to mortality. Increased awareness, comprehensive diagnostic approaches, and targeted therapeutic interventions are critical in managing these cases. Further research is required to elucidate the mechanisms of neuroinflammation in the monkeypox virus and establish standardized treatment protocols.

## INTRODUCTION

Monkeypox virus (MPXV), a zoonotic pathogen belonging to the Orthopoxvirus genus of the Poxviridae family, has garnered significant attention in recent years due to its unprecedented global spread and potential for human-to-human transmission [1,2]. Historically, MPXV has been classified into two distinct genetic clades: the Congo Basin clade (Clade I) and the West African clade (Clade II). Clade I is generally considered more virulent, with a case fatality ratio of 1-10%, compared to Clade II, which has a mortality rate of <3% [3,4].

Since May 2022, a steep increase in MPXV cases has been observed, predominantly in Europe and the United States, marking a significant shift in the epidemiology of this once-rare disease [2,5]. This outbreak, primarily driven by Clade II, has been characterized by novel transmission patterns and clinical presentations, prompting the World Health Organization to declare it a Public Health Emergency of International Concern in July 2022 [6].

While MPXV is primarily known for its characteristic dermatological manifestations, such as rash and skin lesions, emerging evidence suggests that the infection can lead to severe neuro-inflammatory complications [3,7]. These neurological manifestations range from mild symptoms like headache and fatigue to more severe conditions such as encephalitis, meningitis, and seizures, potentially significantly impacting patient outcomes and quality of life [3,8].

The neuro-inflammatory consequences of MPXV infection are poorly understood, and the mechanisms underlying viral invasion and replication within the central nervous system (CNS) remain largely unexplored. The immunological responses triggered by MPXV infection, including cytokine dysregulation and immune cell activation, may contribute to developing neuro-inflammatory manifestations [4,9]. While rare, studies on smallpox infection and vaccination have revealed several cases of encephalitis, seizures, and stroke, providing valuable insights into the neurological complications of orthopoxvirus infections [5,6,10].

Notably, post-vaccination encephalitis following smallpox vaccination has been reported at a rate of 2 to 1219 cases per 100,000 vaccinations, with a higher association observed with neurotrophic vaccinia strains [7,11]. This historical context underscores the importance of understanding orthopoxvirus biology in the etiopathogenesis of neurological sequelae and raises concerns about potential complications as MPXV vaccination strategies are developed and implemented.

The purpose of this systematic review is to summarize evidence documenting de novo MPXV-mediated neuroinflammatory manifestations of the brain and spinal cord. We focus on neuroimmune diseases of the CNS reported in patients who have suffered from MPXV infection rather than in individuals with pre-existing primary demyelinating disorders. This paper aims to provide a comprehensive overview of the current understanding of MPXV infection’s neuro-inflammatory manifestations and outcomes, emphasizing the pathogenesis, clinical manifestations, and immunological correlates of CNS involvement.

Understanding the neuroimmunological perspective of MPXV infection is essential for developing effective therapeutic strategies, and management plans to mitigate the impact of this emerging viral threat. As the global MPXV situation continues to evolve, this review seeks to inform clinicians, researchers, and public health officials about the potential neurological complications of MPXV infection and guide future research directions in this rapidly developing field.

## METHODS

### Design

This systematic review adhered to the Preferred Reporting Items for Systematic Reviews and Meta-Analyses (PRISMA) guidelines (CRD42024583269). We included studies relevant to Monkeypox virus (MPXV) infection cases with suspected or confirmed central nervous system (CNS) inflammatory disorders.

### Search Strategy

We employed pre-specified search strategies to collect data from PubMed, EMBASE, Cochrane Library, Web of Science, and PsycINFO databases up to September 10 2024. The search strategy combined terms indicating MPXV infection with those reflecting CNS inflammatory manifestations. We used relevant Medical Subject Headings (MeSH) and keywords, including ‘Mpox (monkeypox)’, ‘monkeypox virus’, ‘MPV’, ‘Central nervous system diseases’, ‘neurological disorder’, ‘neurologic manifestations’, ‘neurogenic inflammation’, ‘encephalitis’, ‘meningitis’, ‘myelitis’, ‘Miller-Fisher syndrome’, ‘demyelination’, ‘demyelinating autoimmune diseases, CNS’, ‘Spinal Cord Diseases’, ‘transverse myelitis’, ‘multiple sclerosis’, ‘meningoencephalitis’, and ‘encephalomyelitis’.

We also hand-searched additional MPXV-specific articles using reference lists of selected studies, relevant journal websites, and pre-print servers (medRxiv, bioRxiv, and pre-prints.org) from 2022 to the present. To mitigate publication bias, we examined references of all studies potentially missed in the electronic search. Context experts also searched grey literature for relevant articles.

### Study Selection Criteria

We included all peer-reviewed and pre-print (not peer-reviewed) cohort studies, case-control studies, case series, and case reports that met our pre-specified inclusion and exclusion criteria.

#### Inclusion criteria

We included studies that met the following conditions: (i) Studies of MPXV-positive patients with suspected or confirmed neuroinflammatory diseases of the brain or spinal cord (CNS); (ii) MPXV infection-related studies revealing possible associations with neuroimmune disorders or multiple sclerosis with suspected or confirmed brain and spinal cord involvement; (iii) Studies published in English.

Concurrently, we conducted a parallel search to provide a comparative and retrospective outlook on the distribution and extent of cases with similar neurological manifestations in previous outbreaks during 2003–2004 in the United States, 2018–2021 in the United Kingdom, and 2022–2023 in India. We also analyzed the variance of such neurological involvement across different clades of MPXV (Clade I, Clade Ib, and Clade II [IIa, IIb]).

#### Exclusion criteria

We excluded studies in which MPXV was not confirmed and those written in languages other than English. We also excluded review papers, viewpoints, perspectives, commentaries, and studies that did not report information about brain and spinal cord neuroimmune diseases.

### Data Extraction

Prior to screening, three reviewers (GB, VS, and PS) participated in calibration and screening exercises. The first reviewer (GB) independently screened titles and abstracts of all identified citations. The second and third reviewers (VS and PS) verified these citations and screened papers selected by the first reviewer. Another reviewer (RM) independently retrieved and screened full texts of all eligible citations and analyzed the data. Two additional reviewers (SD and JBL) independently verified these full texts for eligibility and designed the overall study structure. JBL resolved disagreements when necessary and made final decisions regarding the study.

Throughout the screening and data extraction process, reviewers used piloted forms. In addition to relevant clinical data, reviewers extracted information on study characteristics (i.e., study identifier, design, setting, and timeframe), outcomes (qualitative or quantitative), clinical factors (definition and measurement methods), and study limitations. The Newcastle-Ottawa Scale was used to assess the selection procedure, comparability, and outcomes of each reviewed study.

### Statistical Analysis

Both quantitative and qualitative data were expressed as percentages. Discordances among variables were resolved by converting them to a standard unit of measurement. A p-value <0.05 was considered statistically significant but could not be calculated due to insufficient data. A meta-analysis was initially planned to analyze associations between demographic findings, symptoms, biochemical parameters, and outcomes but was omitted due to insufficient data.

## RESULTS

We identified 760 articles from databases and 178 from pre-print servers. After removing duplicates, 770 articles were selected. Of these, 650 were excluded after screening titles and abstracts, leaving 120 articles for full-text review. Based on inclusion and exclusion criteria, 82 articles were excluded, along with several others, due to study type (e.g., review papers, correspondence, viewpoints, or commentaries). Thirty-eight articles were finally selected for this study. Of these, 14 articles and one media report were included in the analysis, with the remaining 23 synthesized narratively (**Figure 1, Table 1**).

**Figure 1:**
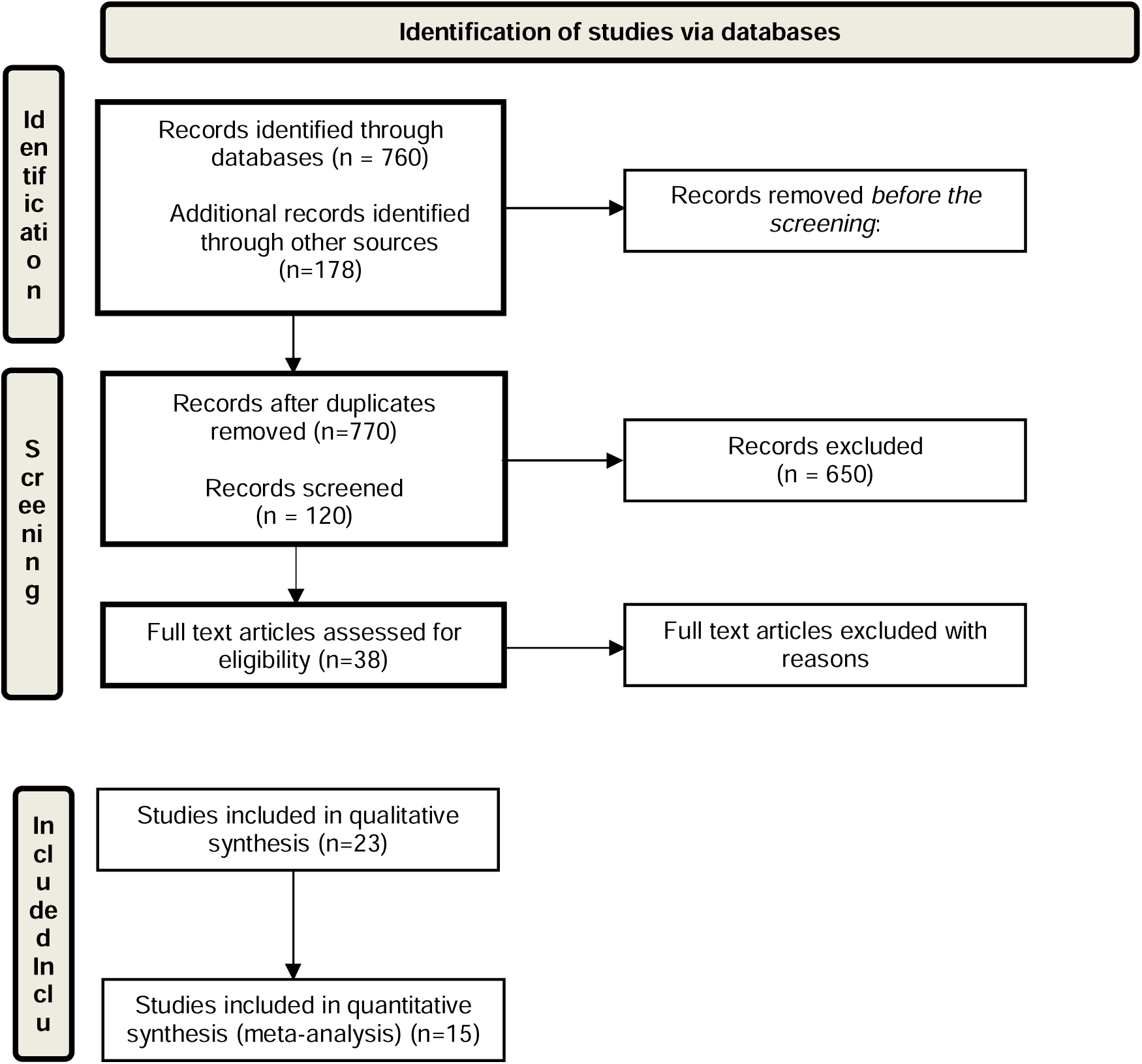
PRISMA flow diagram: identification and selection of studies for the systematic review on monkeypox virus-associated neuroinflammatory disorders.

**Table 1:** Summary of studies reporting monkeypox virus-associated central nervous system neuroinflammatory disorders.

| Authors | Country | Article title | Number of cases |
| --- | --- | --- | --- |
| Hammad et al. [9] | Saudi Arabia | Unusual Neurological Complications in a Patient with Monkeypox: A Case Report | 1 |
| Money et al. [10] | USA | Monkeypox-Associated Central Nervous System Disease: A Case Series and Review | 2 |
| Moore et al. [11] | USA | Transverse Myelitis Associated with Mpox Infection | 1 |
| Cole et al. [12] | UK | Monkeypox Encephalitis with Transverse Myelitis in a Female Patient | 1 |
| Marín-Medina et al. [13] | Colombia | Encephalomyelitis in a Patient with Monkeypox: An Unusual Complication | 1 |
| Pastula et al. [14] | USA | Two Cases of Monkeypox-Associated Encephalomyelitis — Colorado and D.C. | 2 |
| Rodríguez et al. [15] | Colombia | Acute Disseminated Encephalomyelitis in a Patient with Monkeypox: A Case Report | 1 |
| Karin et al. [16] | Sweden | Monkeypox Virus-Associated Meningoencephalitis Diagnosed by Detection of Intrathecal Antibody Production | 1 |
| Yadav et al. [17] | India | An Imported Case of Fatal Encephalitis Associated with Mpox Virus Infection | 1 |
| Sejvar et al. [18] | USA | Human Monkeypox Infection: A Family Cluster in the Midwestern United States | 1 |
| Abdul Hamid et al. [19] | Not reported | Manifestation of Guillain-Barré Syndrome in a Case of Monkeypox Virus Infection | 1 |
| Jezek et al. [20] and [21] | Switzerland | Human Monkeypox: Clinical Features of 282 Patients<br>Clinico-epidemiological features of monkeypox patients with an animal or human source of infection | 2 |
| Ogoina et al. [22] | Nigeria | Clinical Course and Outcome of Human Monkeypox in Nigeria | 3 |
| Reuters (Faus) [23] | Spain | Spain Reports Second Monkeypox-Related Death in Europe | 2 |

**Tables 2 and 3** summarize the demographic and clinical features of patients with central nervous system neuroinflammatory disorders associated with monkeypox virus, and the results of ancillary tests, respectively.

**Table 2:**
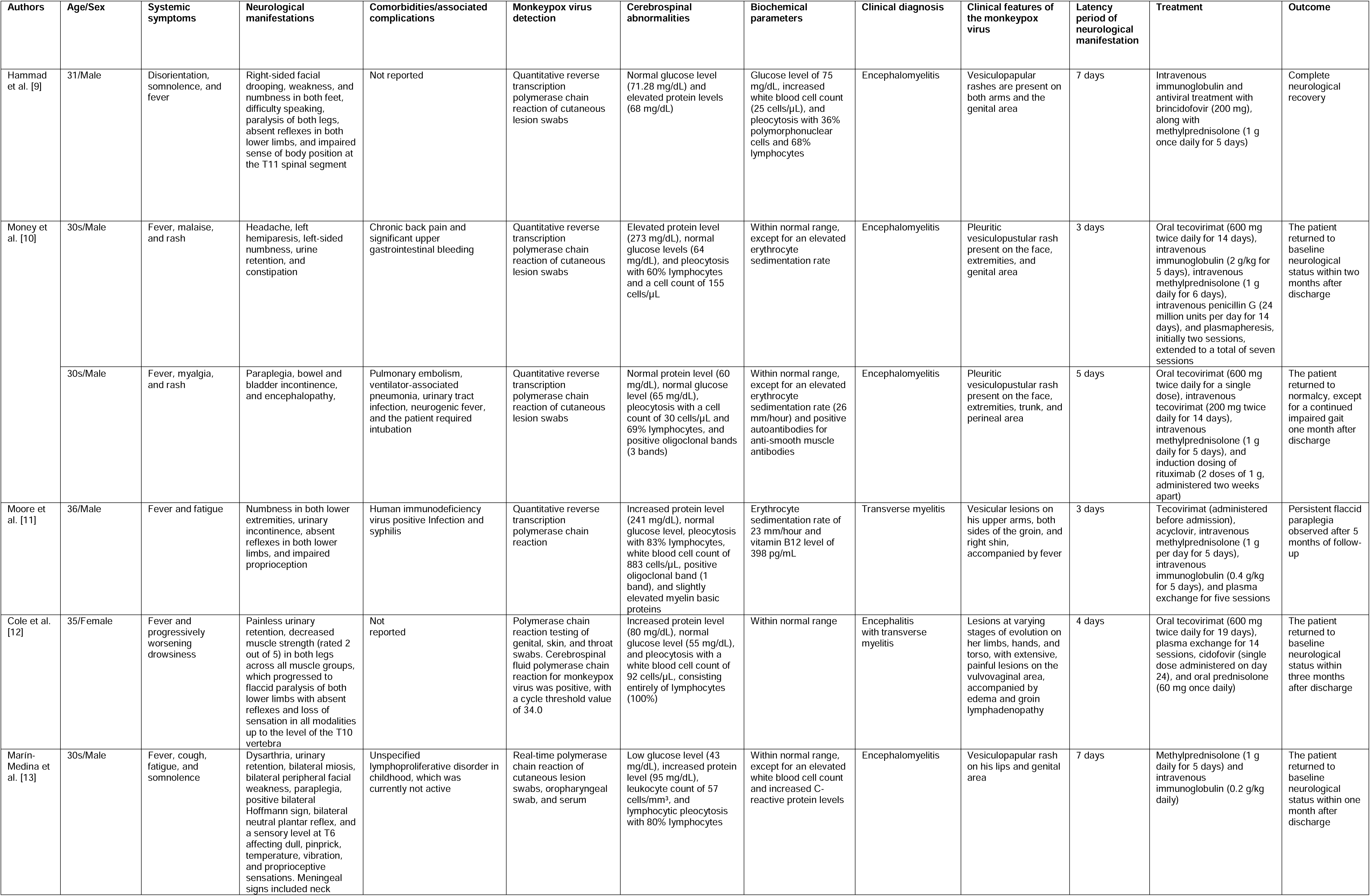
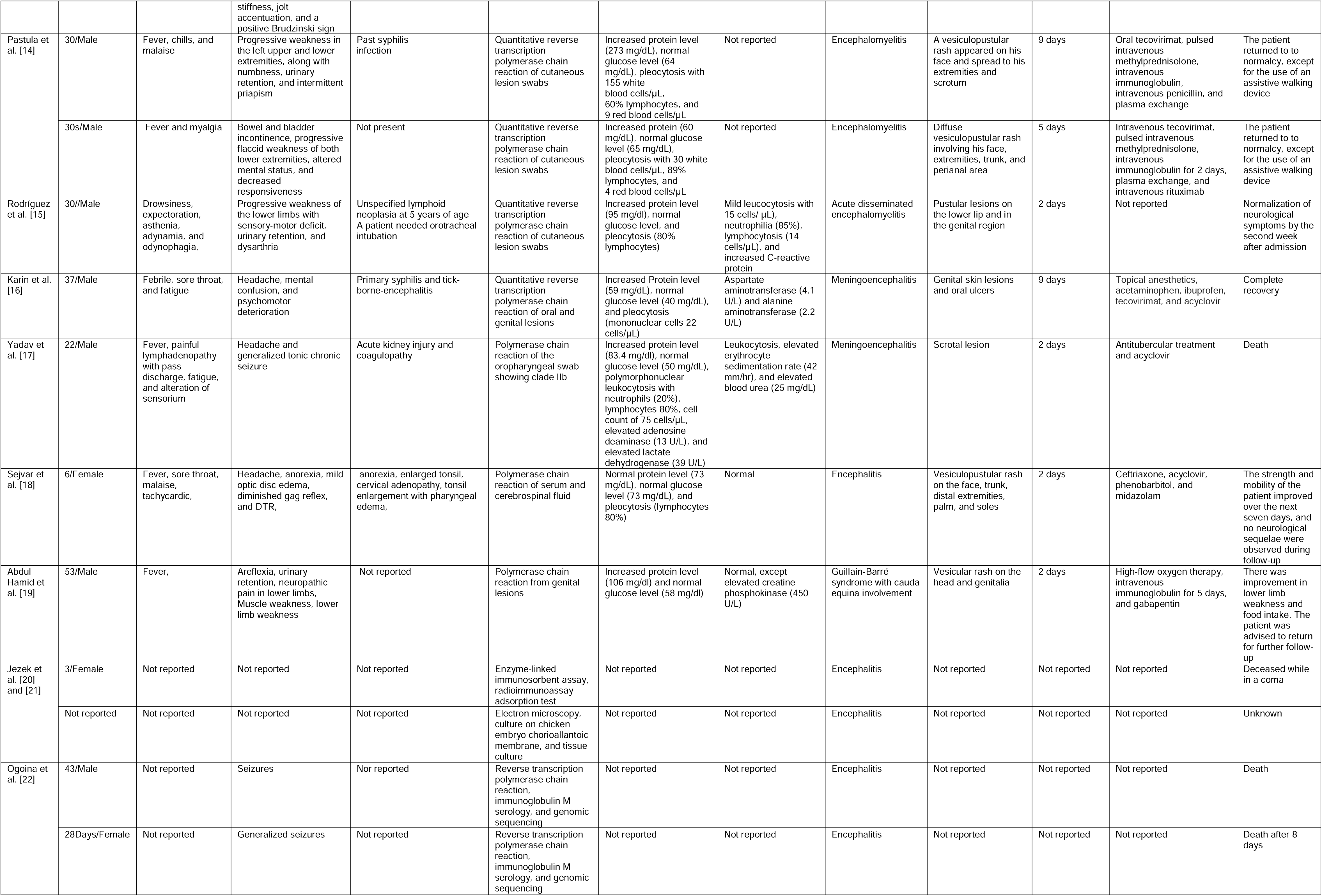
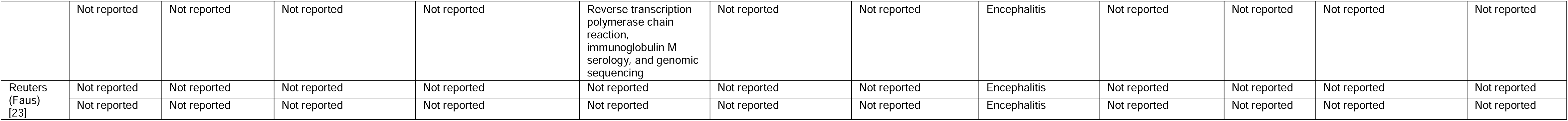
Demographic and clinical features of the patients with monkeypox virus-associated central nervous system neuroinflammatory disorders.

| Authors | Age/Sex | Systemic symptoms | Neurological manifestations | Comorbidities/associated complications | Monkeypox virus detection | Cerebrospinal abnormalities | Biochemical parameters | Clinical diagnosis | Clinical features of the monkeypox virus | Latency period of neurological manifestation | Treatment | Outcome |
| --- | --- | --- | --- | --- | --- | --- | --- | --- | --- | --- | --- | --- |
| Hammad et al. [9] | 31/Male | Disorientation, somnolence, and fever | Right-sided facial drooping, weakness, and numbness in both feet, difficulty speaking, paralysis of both legs, absent reflexes in both lower limbs, and impaired sense of body position at the T11 spinal segment | Not reported | Quantitative reverse transcription polymerase chain reaction of cutaneous lesion swabs | Normal glucose level (71.28 mg/dL) and elevated protein levels (68 mg/dL) | Glucose level of 75 mg/dL, increased white blood cell count (25 cells/μL), and pleocytosis with 36% polymorphonuclear cells and 68% lymphocytes | Encephalomyelitis | Vesiculopapular rashes are present on both arms and the genital area | 7 days | Intravenous immunoglobulin and antiviral treatment with brincidofovir (200 mg), along with methylprednisolone (1 g once daily for 5 days) | Complete neurological recovery |
| Money et al. [10] | 30s/Male | Fever, malaise, and rash | Headache, left hemiparesis, left-sided numbness, urine retention, and constipation | Chronic back pain and significant upper gastrointestinal bleeding | Quantitative reverse transcription polymerase chain reaction of cutaneous lesion swabs | Elevated protein level (273 mg/dL), normal glucose levels (64 mg/dL), and pleocytosis with 60% lymphocytes and a cell count of 155 cells/μL | Within normal range, except for an elevated erythrocyte sedimentation rate | Encephalomyelitis | Pleuritic vesiculopustular rash present on the face, extremities, and genital area | 3 days | Oral tecovirimat (600 mg twice daily for 14 days), intravenous immunoglobulin (2 g/kg for 5 days), intravenous methylprednisolone (1 g daily for 6 days), intravenous penicillin G (24 million units per day for 14 days), and plasmapheresis, initially two sessions, extended to a total of seven sessions | The patient returned to baseline neurological status within two months after discharge |
| | 30s/Male | Fever, myalgia, and rash | Paraplegia, bowel and bladder incontinence, and encephalopathy, | Pulmonary embolism, ventilator-associated pneumonia, urinary tract infection, neurogenic fever, and the patient required intubation | Quantitative reverse transcription polymerase chain reaction of cutaneous lesion swabs | Normal protein level (60 mg/dL), normal glucose level (65 mg/dL), pleocytosis with a cell count of 30 cells/μL and 69% lymphocytes, and positive oligoclonal bands (3 bands) | Within normal range, except for an elevated erythrocyte sedimentation rate (26 mm/hour) and positive autoantibodies for anti-smooth muscle antibodies | Encephalomyelitis | Pleuritic vesiculopustular rash present on the face, extremities, trunk, and perineal area | 5 days | Oral tecovirimat (600 mg twice daily for a single dose), intravenous tecovirimat (200 mg twice daily for 14 days), intravenous methylprednisolone (1 g daily for 5 days), and induction dosing of rituximab (2 doses of 1 g, administered two weeks apart) | The patient returned to normalcy, except for a continued impaired gait one month after discharge |
| Moore et al. [11] | 36/Male | Fever and fatigue | Numbness in both lower extremities, urinary incontinence, absent reflexes in both lower limbs, and impaired proprioception | Human immunodeficiency virus positive Infection and syphilis | Quantitative reverse transcription polymerase chain reaction | Increased protein level (241 mg/dL), normal glucose level, pleocytosis with 83% lymphocytes, white blood cell count of 883 cells/μL, positive oligoclonal band (1 band), and slightly elevated myelin basic proteins | Erythrocyte sedimentation rate of 23 mm/hour and vitamin B12 level of 398 pg/mL | Transverse myelitis | Vesicular lesions on his upper arms, both sides of the groin, and right shin, accompanied by fever | 3 days | Tecovirimat (administered before admission), acyclovir, intravenous methylprednisolone (1 g per day for 5 days), intravenous immunoglobulin (0.4 g/kg for 5 days), and plasma exchange for five sessions | Persistent flaccid paraplegia observed after 5 months of follow-up |
| Cole et al. [12] | 35/Female | Fever and progressively worsening drowsiness | Painless urinary retention, decreased muscle strength (rated 2 out of 5) in both legs across all muscle groups, which progressed to flaccid paralysis of both lower limbs with absent reflexes and loss of sensation in all modalities up to the level of the T10 vertebra | Not reported | Polymerase chain reaction testing of genital, skin, and throat swabs. Cerebrospinal fluid polymerase chain reaction for monkeypox virus was positive, with a cycle threshold value of 34.0 | Increased protein level (80 mg/dL), normal glucose level (55 mg/dL), and pleocytosis with a white blood cell count of 92 cells/μL, consisting entirely of lymphocytes (100%) | Within normal range | Encephalitis with transverse myelitis | Lesions at varying stages of evolution on her limbs, hands, and torso, with extensive, painful lesions on the vulvovaginal area, accompanied by edema and groin lymphadenopathy | 4 days | Oral tecovirimat (600 mg twice daily for 19 days), plasma exchange for 14 sessions, cidofovir (single dose administered on day 24), and oral prednisolone (60 mg once daily) | The patient returned to baseline neurological status within three months after discharge |
| Marin-Medina et al. [13] | 30s/Male | Fever, cough, fatigue, and somnolence | Dysarthria, urinary retention, bilateral miosis, bilateral peripheral facial weakness, paraplegia, positive bilateral Hoffmann sign, bilateral neutral plantar reflex, and a sensory level at T6 affecting dull, pinprick, temperature, vibration, and proprioceptive sensations. Meningeal signs included neck stiffness, jolt accentuation, and a positive Brudzinski sign | Unspecified lymphoproliferative disorder in childhood, which was currently not active | Real-time polymerase chain reaction of cutaneous lesion swabs, oropharyngeal swab, and serum | Low glucose level (43 mg/dL), increased protein level (95 mg/dL), leukocyte count of 57 cells/mm<sup>3</sup>, and lymphocytic pleocytosis with 80% lymphocytes | Within normal range, except for an elevated white blood cell count and increased C-reactive protein levels | Encephalomyelitis | Vesiculopapular rash on his lips and genital area | 7 days | Methylprednisolone (1 g daily for 5 days) and intravenous immunoglobulin (0.2 g/kg daily) | The patient returned to baseline neurological status within one month after discharge |
| Pastula et al. [14] | 30/Male | Fever, chills, and malaise | Progressive weakness in the left upper and lower extremities, along with numbness, urinary retention, and intermittent priapism | Past syphilis infection | Quantitative reverse transcription polymerase chain reaction of cutaneous lesion swabs | Increased protein level (273 mg/dL), normal glucose level (64 mg/dL), pleocytosis with 155 white blood cells/μL, 60% lymphocytes, and 9 red blood cells/μL | Not reported | Encephalomyelitis | A vesiculopustular rash appeared on his face and spread to his extremities and scrotum | 9 days | Oral tecovirimat, pulsed intravenous methylprednisolone, intravenous immunoglobulin, intravenous penicillin, and plasma exchange | The patient returned to to normalcy, except for the use of an assistive walking device |
|  | 30s/Male | Fever and myalgia | Bowel and bladder incontinence, progressive flaccid weakness of both lower extremities, altered mental status, and decreased responsiveness | Not present | Quantitative reverse transcription polymerase chain reaction of cutaneous lesion swabs | Increased protein (60 mg/dL), normal glucose level (65 mg/dL), pleocytosis with 30 white blood cells/μL, 89% lymphocytes, and 4 red blood cells/μL | Not reported | Encephalomyelitis | Diffuse vesiculopustular rash involving his face, extremities, trunk, and perianal area | 5 days | Intravenous tecovirimat, pulsed intravenous methylprednisolone, intravenous immunoglobulin for 2 days, plasma exchange, and intravenous rituximab | The patient returned to to normalcy, except for the use of an assistive walking device |
| Rodríguez et al. [15] | 30//Male | Drowsiness, expectoration, asthenia, adynamia, andodynophagia, | Progressive weakness of the lower limbs with sensory-motor deficit, urinary retention, and dysarthria | Unspecified lymphoid neoplasia at 5 years of age<br>A patient needed orotracheal intubation | Quantitative reverse transcription polymerase chain reaction of cutaneous lesion swabs | Increased protein level (95 mg/dl), normal glucose level, and pleocytosis (80% lymphocytes) | Mild leucocytosis with 15 cells/ μL, neutrophilia (85%), lymphocytosis (14 cells/μL), and increased C-reactive protein | Acute disseminated encephalomyelitis | Pustular lesions on the lower lip and in the genital region | 2 days | Not reported | Normalization of neurological symptoms by the second week after admission |
| Karin et al. [16] | 37/Male | Febrile, sore throat, and fatigue | Headache, mental confusion, and psychomotor deterioration | Primary syphilis and tick-borne-encephalitis | Quantitative reverse transcription polymerase chain reaction of oral and genital lesions | Increased Protein level (59 mg/dL), normal glucose level (40 mg/dL), and pleocytosis (mononuclear cells 22 cells/μL) | Aspartate aminotransferase (4.1 U/L) and alanine aminotransferase (2.2 U/L) | Meningoencephalitis | Genital skin lesions and oral ulcers | 9 days | Topical anesthetics, acetaminophen, ibuprofen, tecovirimat, and acyclovir | Complete recovery |
| Yadav et al. [17] | 22/Male | Fever, painful lymphadenopathy with pass discharge, fatigue, and alteration of sensorium | Headache and generalized tonic chronic seizure | Acute kidney injury and coagulopathy | Polymerase chain reaction of the oropharyngeal swab showing clade IIb | Increased protein level (83.4 mg/dl), normal glucose level (50 mg/dL), polymorphonuclear leukocytosis with neutrophils (20%), lymphocytes 80%, cell count of 75 cells/μL, elevated adenosine deaminase (13 U/L), and elevated lactate dehydrogenase (39 U/L) | Leukocytosis, elevated erythrocyte sedimentation rate (42 mm/hr), and elevated blood urea (25 mg/dL) | Meningoencephalitis | Scrotal lesion | 2 days | Antitubercular treatment and acyclovir | Death |
| Sejvar et al. [18] | 6/Female | Fever, sore throat, malaise, tachycardic, | Headache, anorexia, mild optic disc edema, diminished gag reflex, and DTR, | anorexia, enlarged tonsil, cervical adenopathy, tonsil enlargement with pharyngeal edema, | Polymerase chain reaction of serum and cerebrospinal fluid | Normal protein level (73 mg/dL), normal glucose level (73 mg/dL), and pleocytosis (lymphocytes 80%) | Normal | Encephalitis | Vesiculopustular rash on the face, trunk, distal extremities, palm, and soles | 2 days | Ceftriaxone, acyclovir, phenobarbitol, and midazolam | The strength and mobility of the patient improved over the next seven days, and no neurological sequelae were observed during follow-up |
| Abdul Hamid et al. [19] | 53/Male | Fever, | Areflexia, urinary retention, neuropathic pain in lower limbs, Muscle weakness, lower limb weakness | Not reported | Polymerase chain reaction from genital lesions | Increased protein level (106 mg/dl) and normal glucose level (58 mg/dl) | Normal, except elevated creatine phosphokinase (450 U/L) | Guillain-Barré syndrome with cauda equina involvement | Vesicular rash on the head and genitalia | 2 days | High-flow oxygen therapy, intravenous immunoglobulin for 5 days, and gabapentin | There was improvement in lower limb weakness and food intake. The patient was advised to return for further follow-up |
| Jezek et al. [20] and [21] | 3/Female | Not reported | Not reported | Not reported | Enzyme-linked immunosorbent assay, radioimmunoassay adsorption test | Not reported | Not reported | Encephalitis | Not reported | Not reported | Not reported | Deceased while in a coma |
|  | Not reported | Not reported | Not reported | Not reported | Electron microscopy, culture on chicken embryo chorioallantoic membrane, and tissue culture | Not reported | Not reported | Encephalitis | Not reported | Not reported | Not reported | Unknown |
| Ogoina et al. [22] | 43/Male | Not reported | Seizures | Nor reported | Reverse transcription polymerase chain reaction, immunoglobulin M serology, and genomic sequencing | Not reported | Not reported | Encephalitis | Not reported | Not reported | Not reported | Death |
|  | 28Days/Female | Not reported | Generalized seizures | Not reported | Reverse transcription polymerase chain reaction, immunoglobulin M serology, and genomic sequencing | Not reported | Not reported | Encephalitis | Not reported | Not reported | Not reported | Death after 8 days |
|  | Not reported | Not reported | Not reported | Not reported | Reverse transcription polymerase chain reaction, immunoglobulin M serology, and genomic sequencing | Not reported | Not reported | Encephalitis | Not reported | Not reported | Not reported | Not reported |
| Reuters (Faus) [23] | Not reported | Not reported | Not reported | Not reported | Not reported | Not reported | Not reported | Encephalitis | Not reported | Not reported | Not reported | Not reported |
|  | Not reported | Not reported | Not reported | Not reported | Not reported | Not reported | Not reported | Encephalitis | Not reported | Not reported | Not reported | Not reported |

**Table 3:**
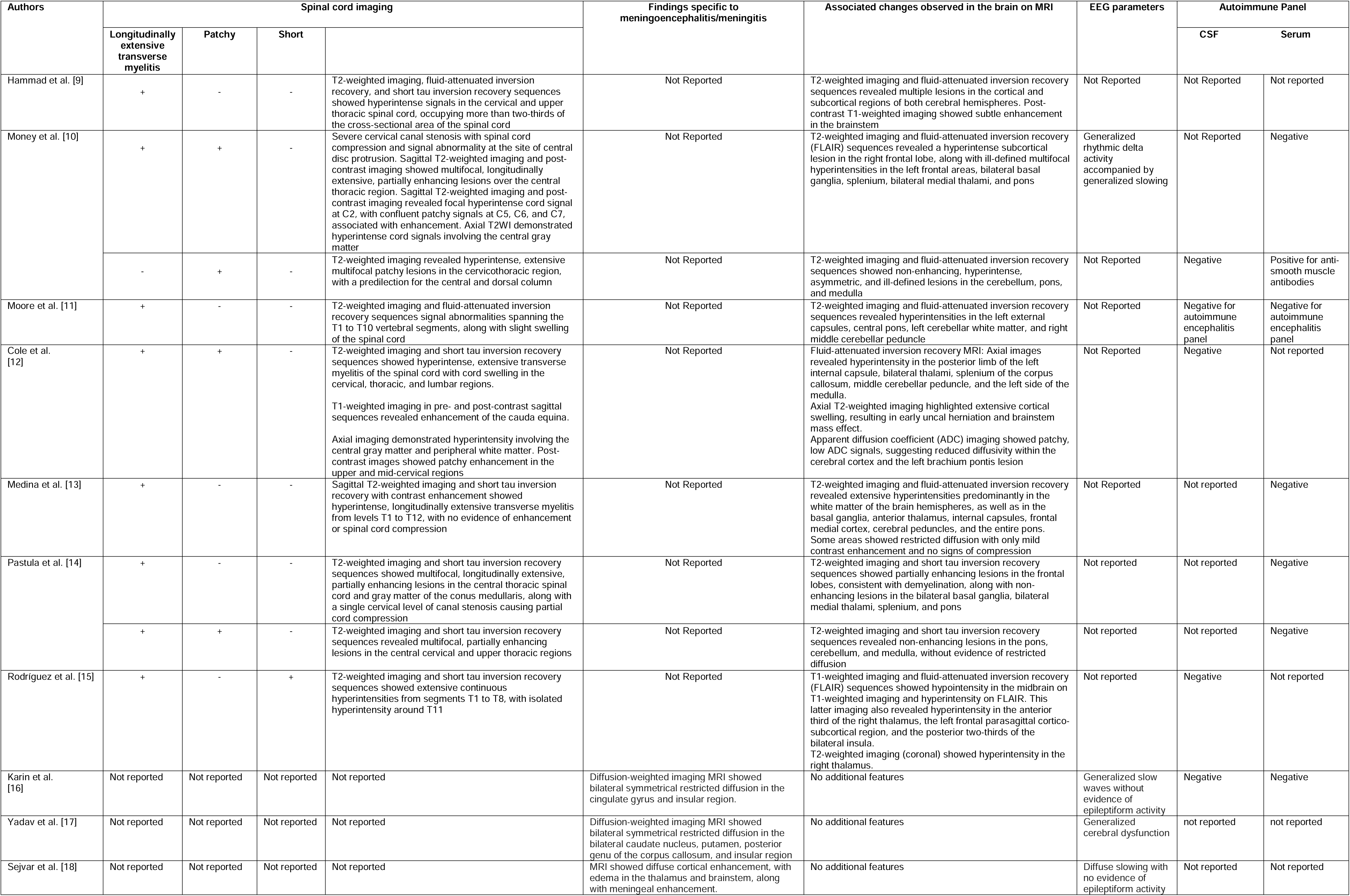
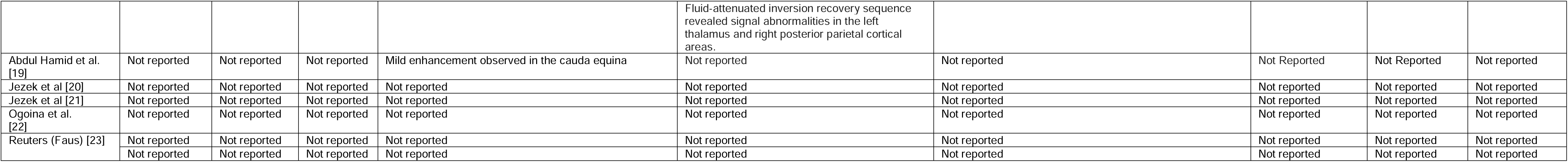
Summary of the studies reporting neuroimaging findings, electroencephalogram, and autoimmune panels associated with MPXV-associated neuroinflammatory disorders.

### Demography and clinical diagnosis

Among the 20 monkeypox-infected cases, 6 were reported from the USA, 3 from Nigeria, 2 each from Spain and Colombia, and 1 each from Saudi Arabia, India, Switzerland, and Sweden (**Figure 2**). Of the 20 cases, 12 were men (75%) and 4 were women (25%), with 4 cases not reporting age and sex. The mean age was 27.9 ± 13.69 years. Diagnoses included encephalitis (8 cases, 40%), encephalomyelitis (6 cases, 30%), meningoencephalitis (2 cases, 10%), isolated transverse myelitis (1 case, 5%), transverse myelitis with encephalitis (1 case, 5%), acute disseminated encephalomyelitis (ADEM) (1 case, 5%), and Guillain–Barré syndrome (GBS) with cauda equina involvement (1 case, 5%).

**Figure 2:**
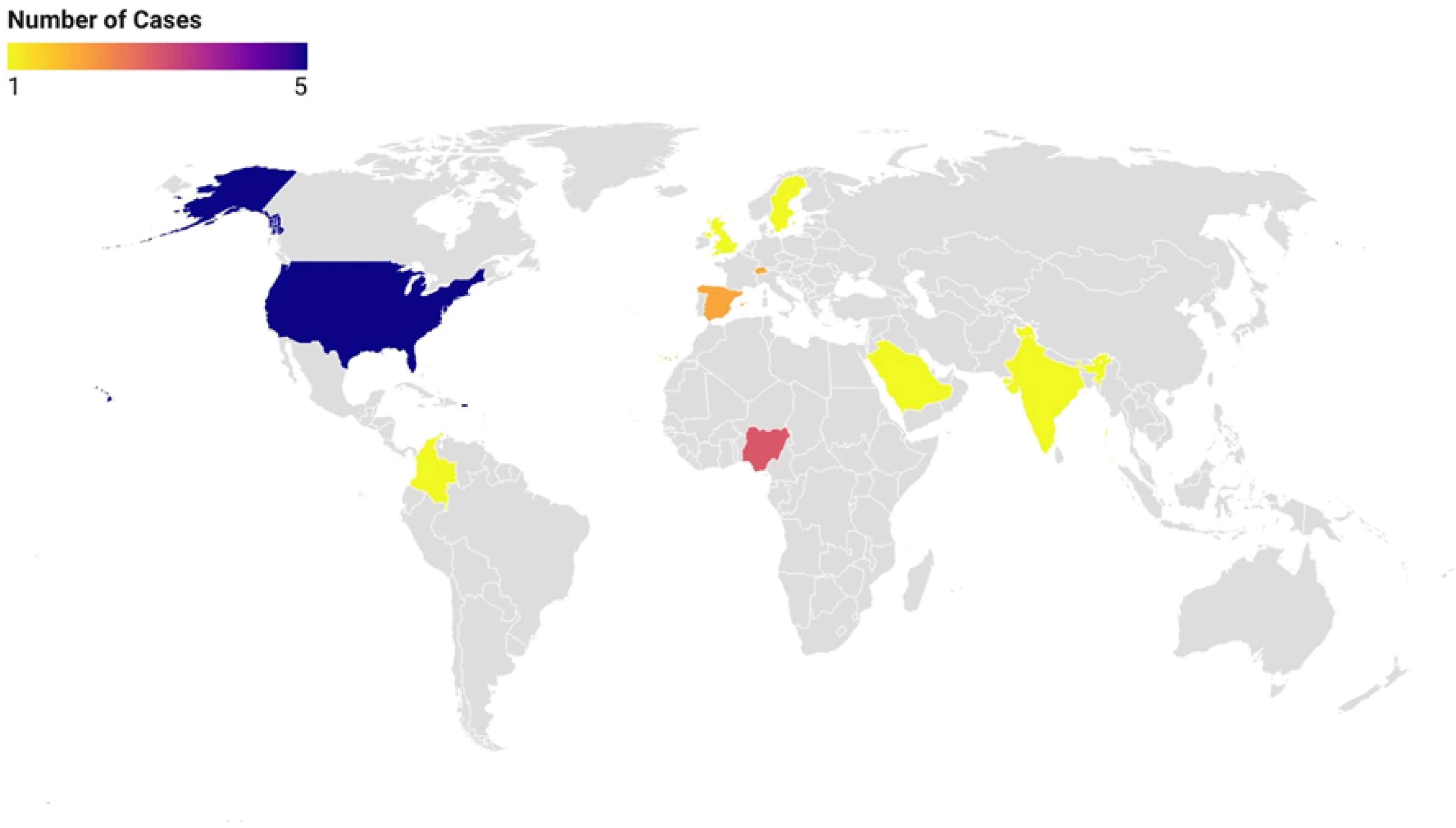
Global distribution of reported cases of neuroinflammatory disorders associated with monkeypox virus.

### Systemic symptoms, co-morbidities, and complications during hospital stay

Systemic symptoms were reported in 13 cases (65%). The most common systemic symptoms were fever (92.3%), fatigue (30.8%), malaise (23.1%), and myalgia (15.4%). Other symptoms included somnolence, sore throat, drowsiness, asthenia, odynophagia, and painful lymphadenopathy.

Associated comorbidities and complications were reported in 13 cases. The most common comorbidities were syphilis (3 cases), lymphoproliferative disorders or lymphoid neoplasia (2 cases), tick-borne encephalitis (1 case), and HIV co-infection (1 case). Reported complications during hospital stay included ventilator-associated pneumonia (1 case) and upper gastrointestinal bleeding (1 case).

### MPXV detection, skin lesion distribution, and characteristics

Of the 18 cases reporting MPXV detection techniques, 16 used quantitative reverse transcription polymerase chain reaction (qRT-PCR) (88.9%), 1 used ELISA (5.6%), 1 used RIA adsorption test (5.6%), and 1 used electron microscopy (5.6%). For qRT-PCR, samples were collected from cutaneous lesion swabs (8 cases), genital lesion swabs (3 cases, 18.8%), oropharyngeal swabs (2 cases, 12.5%), and CSF (2 cases, 12.5%).

MPXV-related skin lesions were reported in 13 cases, with 10 cases (76.9%) associated with defined lesions and 3 cases (23.1%) with undefined characteristics. Among the 10 cases with defined lesions, 5 reported vesiculopustular lesions (50%), 2 reported vesiculopapular lesions (20%), 2 revealed only vesicular features (20%), and 1 was associated with only pustular lesions (10%). One case with undefined lesions showed skin lesions at varying stages.

Regarding anatomical distribution (reported in 13 cases), 8 cases presented with lesions on the extremities (61.5%), of which 6 showed lesions on both upper and lower extremities, and 2 showed lesions only on the upper extremities. Additionally, lesions were found on the face (5 cases, 38.5%), lips (2 cases, 15.4%), trunk (4 cases, 30.8%), genital areas (8 cases, 61.5%), groin (4 cases, 30.8%), perineal area (2 cases, 15.4%), and oral mucosa (1 case, 7.7%).

### Neurological manifestations

The mean latency period for the onset of neurological manifestations was 4.6 ± 2.5 days. Neurological symptoms were available for 13 individuals. The most common manifestations included urinary retention (46.2%), headache (30.8%), altered sensorium and confusion (23.1%), urinary incontinence (23.1%), and seizure (23.1%). Bowel incontinence was reported in 7.7% of cases.

Motor symptoms were reported in 8 cases (61.5%), including weakness in both limbs (50%), left hemiparesis (25%), facial weakness (25%), paraplegia (37.5%), flaccid paralysis (25%), and speech impairment (50%). Common sensorimotor symptoms included areflexia (4 cases) and diminished reflexes (2 cases, 1 DTR, and 1 gag reflex). The most common sensory symptoms were numbness (4 cases), decreased proprioception (3 cases), and decreased pinprick sensation and vibration. One report mentioned complete sensory deficit, and another reported intermittent priapism.

### Biochemical and laboratory parameters

CSF analysis was available for 13 cases, and serum analysis for 11 cases. CSF analysis revealed elevated protein and normal glucose in 10 cases (76.9%), elevated protein and low glucose in 1 case (7.7%), and normal protein and glucose in 2 cases (15.4%). Pleocytosis was observed in 11 cases (84.6%), and oligoclonal bands were found in 2 cases (15.4%) (one case with one band and another with three bands). The CSF autoimmune panel was negative in 5 cases (38.5%).

Serum parameters showed increased leukocytosis and elevated C-reactive protein in 2 cases (18.2%), increased erythrocyte sedimentation rate in 3 cases (27.3%), and lymphocytosis in 7 cases (63.6%). The serum autoimmune panel was reported in 7 cases (63.6%), with only 1 case positive for anti-smooth muscle antibody.

### Electroencephalography results

Electroencephalogram results were available for 4 cases. Generalized slowing without epileptiform activities and patterns indicating generalized cerebral dysfunction were associated with meningoencephalitis. One case of encephalomyelitis showed generalized rhythmic delta activity with generalized slowing, while another case of encephalitis showed diffuse slowing without epileptiform activity [Table 3].

### Neuroimaging findings

Magnetic resonance imaging (MRI) findings were available for 13 cases. Spinal MRI was performed in 10 cases, and brain MRI in 12 cases. Isolated long-extensive transverse myelitis (LETM) was present in 40% of cases, while 3 cases (30%) exhibited coexistence of LETM and patchy involvement, and 1 case (10%) showed both LETM and short segment involvement. Isolated patchy involvement was reported in one patient, and another displayed mild enhancement in the cauda equina. Neuroinflammatory changes in both the brain (including cerebellum and brainstem) and spine were reported in 9 cases (69.2%) [Table 3].

### Treatment and outcome

Treatment modalities were available for 12 patients. The primary antiviral used was tecovirimat (58.3%), administered orally in five cases, intravenously (IV) in one case, and as a combination of oral and IV in one case. Additional antivirals included acyclovir (33.3%) and cidofovir (8.3%). IV penicillin was administered in two cases.

Immunomodulatory therapy consisted of IV methylprednisolone (66.7%), IV immunoglobulins (58.3%), and rituximab (16.7%). Plasmapheresis was utilized in five patients [Table 2].

Seven patients were recommended for follow-up upon discharge. Three demonstrated full recovery, and two regained normal function with the aid of a walking device. Unfortunately, four patients died during the study period.

## DISCUSSION

The emergence of MPXV as a global health concern has brought to light its potential for causing severe neurological complications. This systematic review has identified 20 cases of MPXV-associated neuroinflammatory disorders of the CNS, highlighting the need for increased awareness and understanding of these rare but significant sequelae.

Since the beginning of 2023, the global impact of MPXV has intensified dramatically. More than 27,000 cases have occurred in the Democratic Republic of Congo, resulting in over 1,300 deaths [24]. This sudden rise in the case fatality rate is likely driven by the clade 1b variant. Concurrently, clade II has continued circulating, with more than 1000 cases detected in the USA alone in the first six months of 2024. Notably, the clade 1 or Congo Basin strain exhibits more neurovirulence than the clade 2 or West African variant, which may explain the varying incidence and severity of neurological complications observed across different geographical regions [24].

The geographical distribution of cases in our review spans multiple continents, with the majority reported from the USA (N=6), followed by Nigeria N=3), Spain (N=2), and Colombia (N=2), among others. This global spread underscores the importance of international surveillance and collaboration in managing MPXV outbreaks. The predominance of male patients (75%) in our review aligns with the general trend observed in the 2022 MPXV outbreak, where men who have sex with men were disproportionately affected [2].

The neurological manifestations associated with MPXV infection were diverse, including encephalitis (40%), encephalomyelitis (30%), meningoencephalitis (10%), and rarer presentations such as transverse myelitis and Guillain-Barré syndrome. This spectrum of disorders suggests that MPXV can affect various parts of the CNS, potentially through different pathogenic mechanisms. In the case of neurological manifestations of MPXV, immunological mechanisms play a crucial role in giving rise to diverse clinical presentations affecting both the CNS and peripheral nervous system.

The pathophysiology of MPXV-associated neuroinflammatory disorders remains poorly understood. However, several hypotheses can be proposed based on our findings and current knowledge of viral neuroinvasion. The detection of MPXV in the CSF of some patients suggests that the virus can cross the blood-brain barrier and directly infect CNS tissues [10, 12]. The presence of pleocytosis and elevated protein levels in the CSF of most cases (84.61% and 76.92%, respectively) indicates an inflammatory response within the CNS. This could be due to an overactive immune response to viral antigens or molecular mimicry leading to autoimmune processes [4].

The tendency of MPXV to affect neural tissues, particularly in Clade I variants, may explain the observed neurological complications [3, 7]. Some cases, such as Guillain-Barré syndrome, may represent post-infectious autoimmune phenomena triggered by MPXV infection [19]. The immunological signature observed in MPXV patients, characterized by increased levels of inflammatory cytokines and chemokines [4], may contribute to the development of neuroinflammatory disorders.

The diagnosis of MPXV-associated neuroinflammatory disorders requires a high index of suspicion, especially in the context of an ongoing outbreak. Our review highlights the importance of comprehensive diagnostic approaches, including molecular testing (qRT-PCR), CSF analysis, neuroimaging (MRI), and serological testing. qRT-PCR of various clinical specimens was the primary method for confirming MPXV infection (88.9% of cases) [10, 12, 14]. Consistent findings of elevated protein levels and pleocytosis underscore the value of CSF examination in suspected cases of CNS involvement [10, 12, 14]. MRI plays a crucial role in identifying and characterizing CNS lesions [10, 12, 15], while detection of intrathecal antibody production may be valuable in cases where direct viral detection is challenging [16].

The management of MPXV-associated neuroinflammatory disorders remains challenging due to the lack of specific antiviral therapies. Treatment approaches often included a combination of supportive care, antiviral agents (e.g., tecovirimat), and immunomodulatory therapies. Tecovirimat, an orthopoxvirus VP37 envelope-wrapping protein inhibitor, was used in several cases with varying degrees of success [10, 12, 14]. Corticosteroids and intravenous immunoglobulin were employed in some cases, particularly those with suspected immune-mediated pathology [10, 12, 19]. Management of complications such as seizures, autonomic dysfunction, and respiratory failure was crucial in severe cases [10, 12, 14].

Outcomes were variable, ranging from complete recovery to persistent neurological deficits and, in rare cases, death [10, 12, 14, 22]. The heterogeneity in outcomes underscores the need for early recognition and intervention in suspected cases of MPXV-associated neuroinflammatory disorders.

The emergence of MPXV-associated neuroinflammatory disorders has several implications for public health and clinical practice. Enhanced surveillance for neurological complications in MPXV cases is crucial for understanding the actual burden of these disorders. Healthcare providers should be alert to the possibility of neurological involvement in MPXV patients, particularly those presenting with new-onset neurological symptoms. As vaccination strategies against MPXV are developed and implemented, monitoring for potential neurological adverse events will be critical, given the historical precedent of rare neurological complications associated with smallpox vaccination [5, 7].

This systematic review has several limitations. The small number of reported cases (N=20) limits the generalizability of our findings. Publication bias may have led to an overrepresentation of severe or unusual cases. Additionally, the lack of standardized diagnostic criteria and reporting methods for MPXV-associated neurological complications makes comparing cases challenging. It is important to note that data regarding MPXV-associated neuroinflammatory diseases of the CNS are scarce and underrecognized, which may contribute to the limited number of cases identified in this review.

Future research should focus on prospective cohort studies to determine the true incidence and risk factors for neurological complications in MPXV infection, basic science research to elucidate the mechanisms of MPXV neuroinvasion and neuropathogenesis, clinical trials to evaluate the efficacy of antiviral and immunomodulatory therapies and development of standardized diagnostic and reporting criteria for MPXV-associated neurological complications. Particular attention should be given to understanding the differences in neurovirulence between MPXV clades and their implications for clinical management.

In conclusion, this systematic review highlights the potential for MPXV to cause severe neuroinflammatory disorders of the CNS, with emerging evidence suggesting clade-specific differences in neurovirulence. As the global MPXV situation continues to evolve, with significant outbreaks in regions like the DRC and ongoing transmission worldwide, clinicians and public health officials must remain vigilant for these rare but significant complications. The underrecognition of MPXV-associated neuroinflammatory diseases underscores the need for increased awareness, standardized reporting, and further research. Continued international collaboration is essential to improve our understanding and management of MPXV-associated neurological disorders, particularly in the context of different viral clades and their varying clinical presentations.

## Data Availability

Anonymized data will be provided by Dr. Julian Benito-Leon upon request to any researcher who demonstrates appropriate qualifications.

## Acknowledgments

Julián Benito-León is supported by the National Institutes of Health, Bethesda, MD, USA (NINDS #R01 NS39422) and by the Recovery, Transformation, and Resilience Plan at the Ministry of Science and Innovation (grant TED2021-130174B-C33, NETremor).

## Conflict of Interest Statement

The authors declare that they have no conflicts of interest to disclose.

## Authors Roles

Shramana Deb collaborated on 1) the conception, organization, and execution of the research project, 2) the statistical analysis design, and 3) the writing of the first draft of the manuscript.

Ritwick Mondal collaborated on 1) the conception, organization, and execution of the research project, 2) the statistical analysis design, and 3) the writing of the first draft of the manuscript.

Purbita Sen collaborated on 1) data extraction, 2) data organization, and 3) coordination of literature searches from different databases.

Dipanjan Chowdhury collaborated on 1) data extraction, 2) data organization, and 3) coordination of literature searches from different databases.

Shramana Sarkar collaborated on 1) data extraction, 2) data organization, and 3) coordination of literature searches from different databases.

Granthik Banerjee collaborated on 1) data extraction, 2) data organization, and 3) coordination of literature searches from different databases.

Vramanti Sarkar collaborated on 1) data extraction, 2) data organization, and 3) coordination of literature searches from different databases.

Anjan Chowdhury collaborated on 1) data extraction, 2) data organization, 3) literature search strategy from different databases, and 4) Statistical analysis.

Julián Benito-León collaborated on 1) the conception, organization, and execution of the research project and 2) the writing of the first draft of the manuscript.

## Data availability

Data that has been anonymized will be made available upon request to any researcher who demonstrates appropriate qualifications and will be provided by Dr. Julián Benito-León.

## Notes

### Competing Interest Statement

The authors have declared no competing interest.

### Funding Statement

Nil

